# A national survey of early treatment seeking behavior among those with incident SARS-CoV-2 infection

**DOI:** 10.1101/2022.01.13.22269022

**Authors:** Noah Kojima, Matthew Brobeck, Vladimir Slepnev, Jeffrey D. Klausner

## Abstract

**Background:** Despite effective means to treat SARS-CoV-2 infection, the early treatment seeking behavior of those newly diagnosed with infection is not clear.

**Methods:** We surveyed users of a national SARS-CoV-2 testing company to assess the frequency and correlates of early treatment seeking behavior for a positive test result. We recruited adults (18 years or older) who had tested positive for SARS-CoV-2 by PCR at a large clinical laboratory. To be eligible, individuals had to have a positive test result within 7 days of enrollment. Surveys were anonymous and voluntary. We collected data on demographic characteristics, general health care access and utilization, awareness of treatment for COVID-19, treatment seeking behavior, and treatments received. Descriptive statistics and odds ratios (OR) with 95% confidence intervals (95% CI) were calculated on StataSE.

**Results:** Participants were surveyed from 3-7 January 2022: among the 15,991 who viewed a survey request, 7,647 individuals were eligible and provided responses. The median age of a respondent was 42 years (interquartile range: 32 to 54), 68.9% of respondents were women, and respondents represented 33 different states, districts, and territories. Among respondents, 23.1% reported they had sought treatment or medical advice for their current COVID-19 diagnosis. Of those who were very aware of treatment for COVID-19, 31.0% sought treatment versus 16.7% who were unaware (p-value< 0.001). The odds of treatment seeking behavior were higher for those that were contacted by a medical professional after their diagnosis (OR: 4.57 [95% CI: 3.89 to 5.37]), those with a primary doctor (OR: 2.94 [95% CI: 2.52 to 3.43]), those who self-measured their oxygen saturation (OR: 2.53 [95% CI: 2.25 to 2.84]), and those over 65 years of age (OR: 2.36 [95% CI: 2.02 to 2.76]). There was no difference in those seeking treatment based on heritage, ethnicity, prior COVID-19 diagnosis, state political affiliation, or vaccination status. The odds of seeking treatment were lower among men (OR: 0.88 [95% CI: 0.78 to 0.99]) and those without insurance (OR: 0.62 [95% CI: 0.52 to 0.72]). The most common treatment locations were clinics and most common treatments were Vitamin C, Vitamin D, Zinc, Tylenol, and NSAIDs.

**Conclusion:** More public outreach is needed to raise awareness of the benefits of treatment for COVID-19. We found that people who were more aware about treatment for COVID-19 were more likely to seek medical advice or therapy. Efforts to increase awareness might increase early treatment for SARS-CoV-2 infection. Increased outreach with treatment facilitation from medical professionals and/or public health staff to those with newly detected SARS-CoV-2 infections, particularly among those at higher-risk of complications, might also be helpful.

## Main Text

COVID-19 continues to cause severe disease, hospitalization, and death. Despite effective means to treat SARS-CoV-2 infection,^1-4^ the early treatment seeking behavior of those newly diagnosed with infection is not clear. We surveyed users of a national SARS-CoV-2 testing company to assess the frequency and correlates of early treatment seeking behavior for a positive test result.

We recruited adults (18 years or older) who had tested positive for SARS-CoV-2 by PCR at a large clinical laboratory (Curative, San Dimas, CA). To be eligible, individuals had to have a positive test result within 7 days of enrollment. Surveys were anonymous and voluntary. We collected data on demographic characteristics, general health care access and utilization, awareness of treatment for COVID-19, treatment seeking behavior, and treatments received. Descriptive statistics and odds ratios (OR) with 95% confidence intervals (95% CI) were calculated on StataSE (StataCorp, College Station, TX). Chi-squared tests and linear regression analysis were used to compare categorical variables and continuous variables between groups, respectively. Advarra granted the study IRB exempt (Pro00059961).

Participants were surveyed from 3-7 January 2022: among the 15,991 who viewed a survey request, 7,647 individuals were eligible and provided responses. The median age of a respondent was 42 years (interquartile range: 32 to 54), 68.9% of respondents were women, and respondents represented 33 different states, districts, and territories. Among those surveyed, 66.7% identified as White, 30.6% as Hispanic, 12.1% as Black or African American, 6.2% as Asian, and 1.2% as Pacific Islander or Native American. Most respondents reported they were vaccinated (89.3%).

Among respondents, 23.1% reported they had sought treatment or medical advice for their current COVID-19 diagnosis (Table 1). Of those who were very aware of treatment for COVID-19, 31.0% sought treatment versus 16.7% who were unaware (p-value< 0.001). The odds of treatment seeking behavior were higher for those that were contacted by a medical professional after their diagnosis (OR: 4.57 [95% CI: 3.89 to 5.37]), those with a primary doctor (OR: 2.94 [95% CI: 2.52 to 3.43]), those who self-measured their oxygen saturation (OR: 2.53 [95% CI: 2.25 to 2.84]), and those over 65 years of age (OR: 2.36 [95% CI: 2.02 to 2.76]).

**Table 1.**
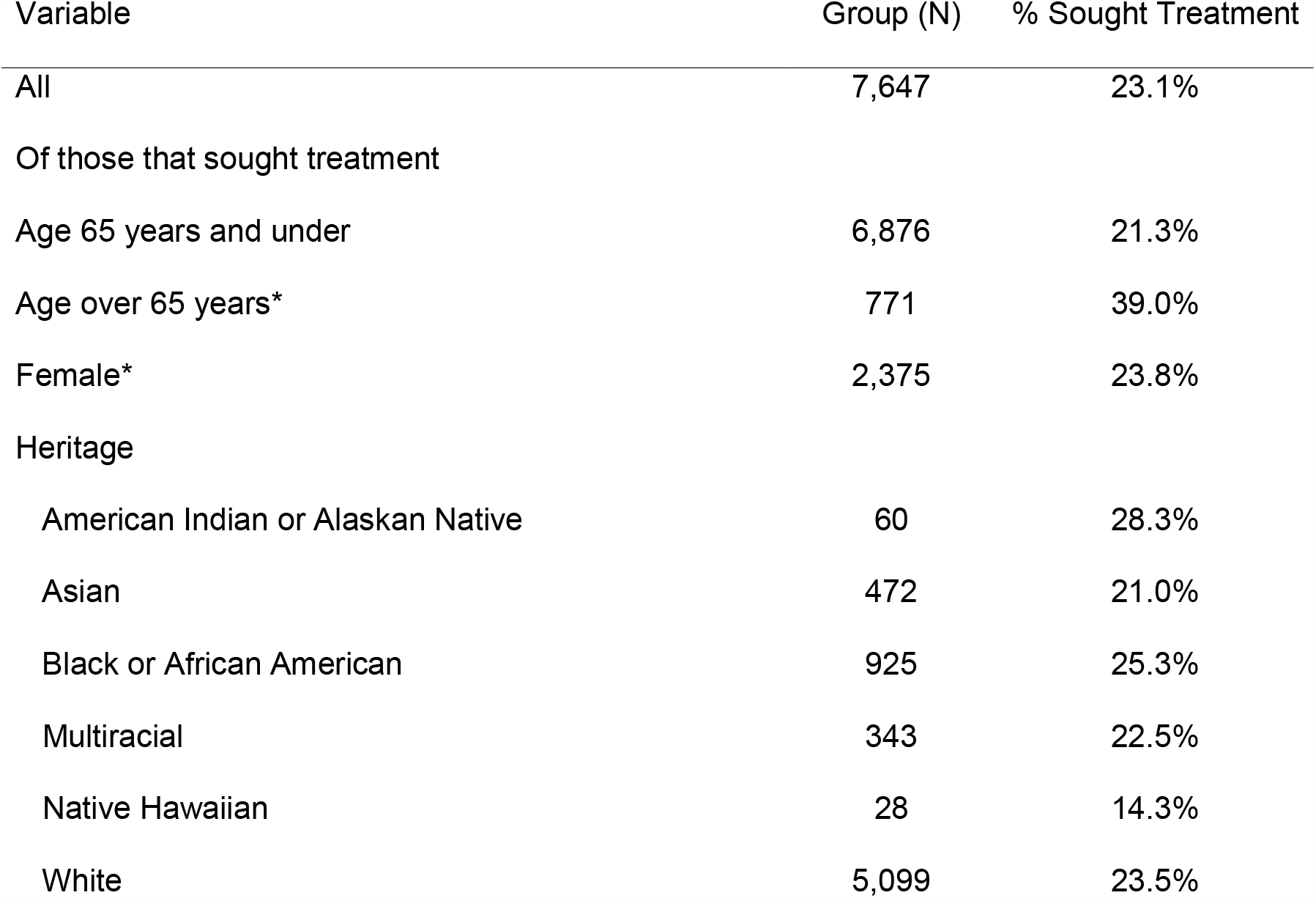
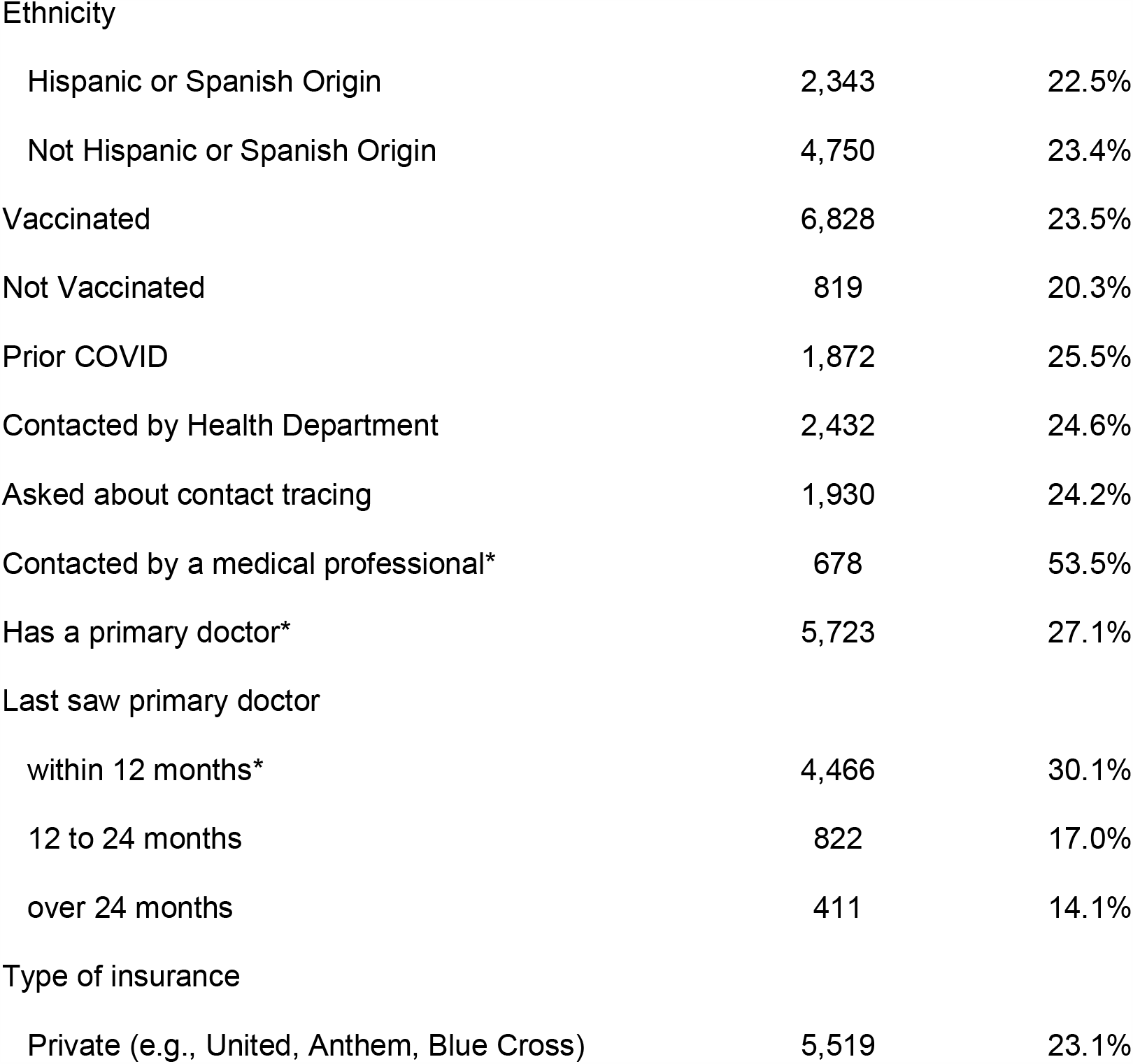
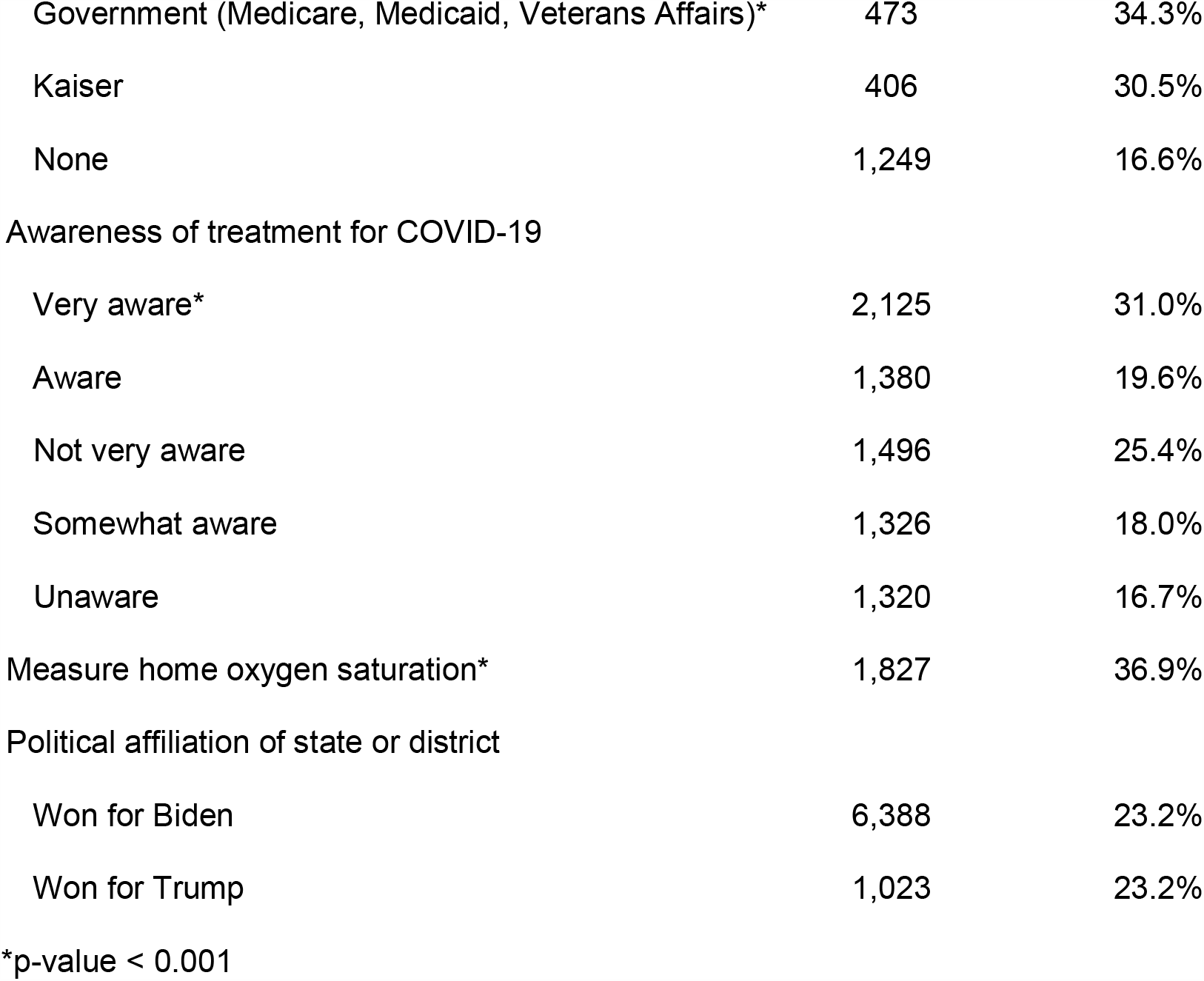
Demographic characteristics, health care access and utilization, awareness of treatment and treatment seeking behavior for COVID-19 among a national cohort of survey respondents that recently tested positive for SARS-CoV-2 infection, 3 January 2022 to 7 January 2022

| Variable | Group (N) | % Sought Treatment |
| --- | --- | --- |
| All | 7,647 | 23.1% |
| Of those that sought treatment |  |  |
| Age 65 years and under | 6,876 | 21.3% |
| Age over 65 years* | 771 | 39.0% |
| Female* | 2,375 | 23.8% |
| Heritage |  |  |
| American Indian or Alaskan Native | 60 | 28.3% |
| Asian | 472 | 21.0% |
| Black or African American | 925 | 25.3% |
| Multiracial | 343 | 22.5% |
| Native Hawaiian | 28 | 14.3% |
| White | 5,099 | 23.5% |
| Ethnicity |  |  |
| Hispanic or Spanish Origin | 2,343 | 22.5% |
| Not Hispanic or Spanish Origin | 4,750 | 23.4% |
| Vaccinated | 6,828 | 23.5% |
| Not Vaccinated | 819 | 20.3% |
| Prior COVID | 1,872 | 25.5% |
| Contacted by Health Department | 2,432 | 24.6% |
| Asked about contact tracing | 1,930 | 24.2% |
| Contacted by a medical professional* | 678 | 53.5% |
| Has a primary doctor* | 5,723 | 27.1% |
| Last saw primary doctor |  |  |
| within 12 months* | 4,466 | 30.1% |
| 12 to 24 months | 822 | 17.0% |
| over 24 months | 411 | 14.1% |
| Type of insurance |  |  |
| Private (e.g., United, Anthem, Blue Cross) | 5,519 | 23.1% |
| Government (Medicare, Medicaid, Veterans Affairs)* | 473 | 34.3% |
| Kaiser | 406 | 30.5% |
| None | 1,249 | 16.6% |
| Awareness of treatment for COVID-19 |  |  |
| Very aware* | 2,125 | 31.0% |
| Aware | 1,380 | 19.6% |
| Not very aware | 1,496 | 25.4% |
| Somewhat aware | 1,326 | 18.0% |
| Unaware | 1,320 | 16.7% |
| Measure home oxygen saturation* | 1,827 | 36.9% |
| Political affiliation of state or district |  |  |
| Won for Biden | 6,388 | 23.2% |
| Won for Trump | 1,023 | 23.2% |

\*p-value < 0.001

There was no difference in those seeking treatment based on heritage, ethnicity, prior COVID-19 diagnosis, state political affiliation, or vaccination status. The odds of seeking treatment were lower among men (OR: 0.88 [95% CI: 0.78 to 0.99]) and those without insurance (OR: 0.62 [95% CI: 0.52 to 0.72]). The most common treatment locations were clinics and most common treatments were Vitamin C, Vitamin D, Zinc, Tylenol, and NSAIDs (Table 2).

**Table 2.** Treatment locations and treatments used for COVID-19 among survey respondents who tested positive for SARS-CoV-2 infection in the prior 7 days, among 1767 persons reporting seeking treatment, 3 January 2022 to 7 January

| Variable | Number | % |
| --- | --- | --- |
| Location of treatment or medical advice |  |  |
| Clinic | 855 | 48.4% |
| Urgent Care | 157 | 8.9% |
| Emergency Department | 88 | 5.0% |
| Mobile Clinic | 71 | 4.0% |
| Free Clinic | 28 | 1.6% |
| Treatment used |  |  |
| Vitamin C | 825 | 46.7% |
| Vitamin D | 685 | 38.8% |
| Zinc | 636 | 36.0% |
| Acetaminophen (Tylenol) | 546 | 30.9% |
| NSAIDs (e.g., Advil, Aleve, Motrin, or Ibuprofen) | 522 | 29.5% |
| Azithromycin (Zithromax) | 203 | 11.5% |
| Dexamethasone (Decadron) or prednisone* | 179 | 10.1% |
| Aspirin | 169 | 9.6% |
| Monoclonal antibodies** | 100 | 5.7% |
| Ivermectin (Stromectol) | 50 | 2.8% |
| Hydroxychloroquine (Plaquenil) | 21 | 1.2% |
| Oxygen | 14 | 0.8% |
| Nirmatrelvir/Ritonavir (Paxlovid; Pfizer) | 13 | 0.7% |
| Fluvoxamine (Luvox, Faverin, Fluvoxin) | 13 | 0.7% |
| Remdesivir (Veklury; Gilead) | 10 | 0.6% |
| Convalescent plasma | 2 | 0.1% |
| Colchicine (Colcrys) | 2 | 0.1% |
| Molnupiravir (Lagevrio; Merck) | 1 | 0.1% |

\*(Deltasone, Mericorten, Liquid Pred, Orasone) or inhaled budesonide (Pulmicort, Duoresp)
\*\*(casiriviman-imdevimab [REGEN-COV]; REGENERON, sotrovimab [Xevudy]; GlaxoSmithKline, or bamlanivimab-
etesevimab; Eli Lilly)

Due to the timing of the survey, infection was most likely due to the Omicron variant of SARS-CoV-2, which might be less severe in those with prior immunity.^5^ The most commonly used treatments were over-the-counter medications for symptom relief. Few had received therapies known to reduce risk of disease progression.^4,6^ There were also continued reports of non-beneficial medication use, i.e., ivermectin and hydroxychloroquine.

More public outreach is needed to raise awareness of the benefits of treatment for COVID-19. We found that people who were more aware about treatment for COVID-19 were more likely to seek medical advice or therapy. Efforts to increase awareness might increase early treatment for SARS-CoV-2 infection. Additionally, those who were contacted by a health professional were more likely to seek treatment or medical advice for COVID-19. Increased outreach with treatment facilitation from medical professionals and/or public health staff to those with newly detected SARS-CoV-2 infections, particularly among those at higher-risk of complications, might also be helpful.

## Data Availability

All data produced in the present study are available upon reasonable request to the authors.

## Acknowledgements

To the participants that donated their time.

